# Characteristics of ictal thalamic EEG in pediatric-onset neocortical focal epilepsy

**DOI:** 10.1101/2023.06.22.23291714

**Authors:** Benjamin Edmonds, Makoto Miyakoshi, Luigi Gianmaria Remore, Samuel Ahn, H. Westley Phillips, Atsuro Daida, Noriko Salamon, Ausaf Bari, Raman Sankar, Joyce H. Matsumoto, Aria Fallah, Hiroki Nariai

**Affiliations:** Division of Pediatric Neurology, Department of Pediatrics, UCLA Mattel Children’s Hospital, David Geffen School of Medicine, Los Angeles, CA, USA; Division of Child and Adolescent Psychiatry, Cincinnati Children’s Hospital Medical Center, Cincinnati, OH, USA; Department of Psychiatry, University of Cincinnati College of Medicine, Cincinnati, OH, USA; Swartz Center for Computational Neuroscience, Institute for Neural Computation, University of California San Diego, UCSD Medical Center, San Diego, CA, USA; Department of Neurosurgery, UCLA Medical Center, David Geffen School of Medicine, Los Angeles, CA, USA; Department of Radiological Sciences, University of California, Los Angeles, CA, USA; The UCLA Children’s Discovery and Innovation Institute, Los Angeles, CA, USA

**Author notes:** Corresponding author: Benjamin Edmonds, MD Division of Pediatric Neurology, Department of Pediatrics, UCLA Mattel Children’s Hospital, David Geffen School of Medicine, Los Angeles, CA, USA. Address: 10833 Le Conte Ave, Room 22-474, Los Angeles, CA 90095-1752, USA.

**Keywords:** neurostimulation, thalamus, drug-resistant epilepsy, epilepsy surgery, RNS

## Abstract

**Objective:** To characterize ictal EEG change in the centromedian (CM) and anterior nucleus (AN) of the thalamus, using stereoelectroencephalography (SEEG) recordings

**Methods:** Forty habitual seizures were analyzed in nine patients with pediatric-onset neocortical drug-resistant epilepsy who underwent SEEG (age 2-25 y) with thalamic coverage. Both visual and quantitative analysis was used to evaluate ictal EEG signal in the cortex and thalamus. The amplitude and cortico-thalamic latencies of broadband frequencies at ictal onset were measured.

**Results:** Visual analysis demonstrated consistent detection of ictal EEG changes in both the CM nucleus and AN nucleus with latency to thalamic ictal EEG changes of less than 400ms in 95% of seizures, with low-voltage fast activity being the most common ictal pattern. Quantitative broadband amplitude analysis showed consistent power changes across the frequency bands, corresponding to ictal EEG onset, while while ictal EEG latency was variable from -18.0 seconds to 13.2 seconds. There was no significant difference between detection of CM and AN ictal activity on visual or amplitude analysis. Four patients with subsequent thalamic responsive neurostimulation (RNS) demonstrated ictal EEG changes consistent with SEEG findings.

**Conclusions:** Ictal EEG changes were consistently seen at the CM and AN of the thalamus during neocortical seizures.

**Significance:** It may be feasible to use a closed-loop system in the thalamus to detect and modulate seizure activity for neocortical epilepsy.

## 1) INTRODUCTION

Drug-resistant epilepsy (DRE) is prevalent in up to 30% of patients with epilepsy (Kwan and Brodie, 2000, Sultana et al., 2021) and leads to an increased risk of mortality and neurodevelopmental comorbidities (Chen et al., 2018, Kwan and Brodie, 2000). Currently, seizure-freedom rates following epilepsy surgery in children remain suboptimal, ranging from 50 to 75% (Asano et al., 2009, Dwivedi et al., 2017, Harris et al., 2022, Hoppe et al., 2022). However, for the substantial subset of patients who have contraindications to resection, including multifocal epilepsy, seizures originating in the eloquent cortex, or significant comorbidities for surgery, neurostimulation provides an alternative and potentially highly effective means of seizure control (Klinger and Mittal, 2018).

At present, there are three main options for neurostimulation: Vagus Nerve Stimulation (VNS) (Gonzalez et al., 2019, Kotagal, 2011), Deep Brain Stimulation (DBS) (Fisher et al., 2010, Salanova et al., 2021, Zangiabadi et al., 2019), and Responsive Neurostimulation (RNS)(Kwon et al., 2020a, Nair et al., 2020). VNS is a well-established open-loop device, approved by FDA in 1997, to treat DRE, and further expanded to pediatrics (≥4 years) in 2017, that delivers a pre-scheduled stimulation, regardless of brain activity, that can be effective in reducing seizure burden by 50% or more (Englot et al., 2016). DBS, like VNS, is an open-loop device, but with the advantage that it can be targeted intracranially to stimulate specific subcortical nuclei and has been FDA approved for adults since 2018 for the treatment of focal or focal with secondary generalization epilepsy via stimulation of the anterior nucleus of the thalamus (AN) (Salanova et al., 2021). Lastly, RNS, FDA approved in 2013 for focal DRE epilepsy, is an attractive option for both adult and pediatric patients given its novel advantage of being a closed-loop system that can record live electrocorticography (ECOG) and stimulate based on a specific electrographic pattern in up to two regions currently(Morrell and Group, 2011, Nagahama et al., 2021). All of these modalities have shown increased effectiveness over time without a clear indication of which is most effective (Ryvlin and Jehi, 2022).

Therefore, the most effective neurostimulation strategy for patients with multifocal or generalized epilepsy with no clear target or too many targets for neurostimulation remains an open question. Recent case reports and small studies have targeted the thalamus, including the AN (Fisher et al., 2010, Herlopian et al., 2019), CM (Burdette et al., 2020, Dalic et al., 2022, Li and Cook, 2018), and pulvinar (Burdette et al., 2021) nucleus, to disrupt seizure activity. While initial studies show potential effectiveness of neurostimulation at the AN, CM, and pulvinar (Beaudreault et al., 2022, Kokkinos et al., 2020, Kwon et al., 2020b, Welch et al., 2021) it has not been determined how sensitive they are at detecting the neocortical seizure onset, i.e. seizure onset pattern (SOP). Moreover, there is no published comparison, qualitative or quantitative, between AN recording and CM recordings to determine which provides a more sensitive signal for ictal activity.

In this study, we aim to evaluate the stereoelectroencephalography (SEEG) ictal signature of the AN and CM of the thalamus in pediatric-onset DRE to determine if there is a clear SOP of habitual seizures in the AN and CM that could be targeted with neurostimulation to treat focal neocortical epilepsy. Moreover, we also aim to determine which thalamic nucleus is more sensitive in detecting the ictal onset, and if a quantitative analysis can detect ictal EEG changes in the thalamus to determine the timing of seizure onset. If thalamic recording (AN or CM) can reliably detect seizure onset, the idea of thalamic RNS for seizure control becomes realistic, enabling a guide for closed-loop neuromodulation, which has the potential to be better than DBS or VNS (open-loop).

## 2) MATERIALS & METHODS

This is a retrospective single-center study conducted at the University of California Los Angeles. The institutional review board (IRB#18-001599) at UCLA approved the use of human subjects and waived the need for written informed consent, as all testing was deemed clinically relevant for patient care. This study was not a clinical trial, and it was not registered in any public registry.

### 2.1) Patient Selection

All patients with neocortical pediatric-onset epilepsy admitted from November 2020 to April 2022 with DRE evaluated by the pediatric epilepsy group at UCLA Mattel Children’s Hospital and underwent a chronic SEEG implantation with electrodes inserted into the thalamus (AN and CM). Those patients who are suspected to have drug resistant generalized epilepsies or multi-focal epilepsies. There were no exclusion criteria.

### 2.2) Study Protocol

The plan for SEEG placement was discussed at our multidisciplinary epilepsy surgery conference (consisting of epileptologists, neurosurgeons, radiologists, and neuropsychologists) and was based on the combination of data from seizure semiology, neurological examination, neuroimaging findings (MRI, PET, and magnetoencephalography), neuropsychological evaluation, and scalp EEG with emphasis primarily on seizure onset zones (Nariai et al., 2019). Ipsilateral (to the presumed site of the presumed seizure onset) CM and AN thalamic SEEG electrodes were placed to determine whether an ictal pattern can be detected in the thalamus and provide a potential target for therapy. If patient had bilateral seizure onsets, thalamic SEEG electrodes were placed on the ipsilateral side of greatest seizure burden, or if unclear, bilateral thalamic electrodes were placed.

#### 2.2.1) SEEG Placement

BrainLab elements software was used for planning the electrodes to the intended targets using T1-weighted sequences, and the trajectories were guided by a gadolinium-enhanced T1-weighted MRI. Target subcortical structures (including the CM and AN thalamic nuclei) were identified and outlined by experienced neuroradiologist (NS) on a case-by-case basis prior to the trajectory planning. The targets and trajectories that were planned using the MRI MPRAGE2 sequence were then co-registered to a volumetric CT scan acquired after placing the patient’s head into the Leksell frame. Using the Leksell coordinates obtained from the BrainLab elements software, each electrode was placed. Four contact Spencer Depth Electrodes with 5 mm spacing were used exclusively for thalamic targets. Intraoperative or immediate postoperative CT scan was used to rule out intracranial hemorrhage and confirm the final position and trajectory of each electrode placed.

#### 2.2.2) Intracranial EEG recording

EEG recording was obtained using Nihon Kohden (Irvine, California, USA). The study recording was acquired with a digital sampling frequency of 200 Hz. This was then reviewed digitally at default proprietary Nihon Kohden settings of a low frequency filter (LFF) of 0.016 Hz and a high frequency filter (HFF) of 70 Hz. All ECoGs were part of the clinical EEG recording (Nariai et al., 2019).

#### 2.2.3) Leads reconstruction

SEEG electrodes were localized using the advanced processing pipeline in Lead-DBS software (Ver 2.6), as previously described elsewhere(Horn et al., 2019). Briefly, postoperative CT was linearly co-registered to pre-operative MRI using advanced normalization tools (ANTs3), brain shift correction was applied and all preoperative volumes were non-linearly co-registered to the MNI (Montreal Neurologic Institute) ICBM 2009b NLIN asymmetric space(Fonov et al., 2011) employing the ANTs SyN Diffeomorphic Mapping. DBS contacts were automatically pre-reconstructed using the phantom-validated and fully automated PaCER method(Husch et al., 2018) or the TRAC/CORE approach and manually refined when appropriate. The resultant electrode models were then wrapped in the MNI space. Atlas segmentations in this manuscript are defined by the THOMAS atlas to visualize ANT and CM(Su et al., 2019). Group visualization and analysis of active contact location were performed using the Lead-Group tool in Lead-DBS(Horn et al., 2019).

### 2.3) Confirmation of Electrode Placement

The location of electrode placement was verified post-operatively with CT co-registered with the pre-op T1-weighted MRI using the BrainLab Elements software. Outlines for target subcortical nuclei were identified and overlayed onto the postoperative CT scan. Electrode contact placement in relation to the target nuclei was then determined. For contacts placed outside of the desired nucleus, distance-to-target measurements were then recorded from the edge of the respective contact to the edge of the nucleus of interest (**Figure 1**).

**Figure 1:**
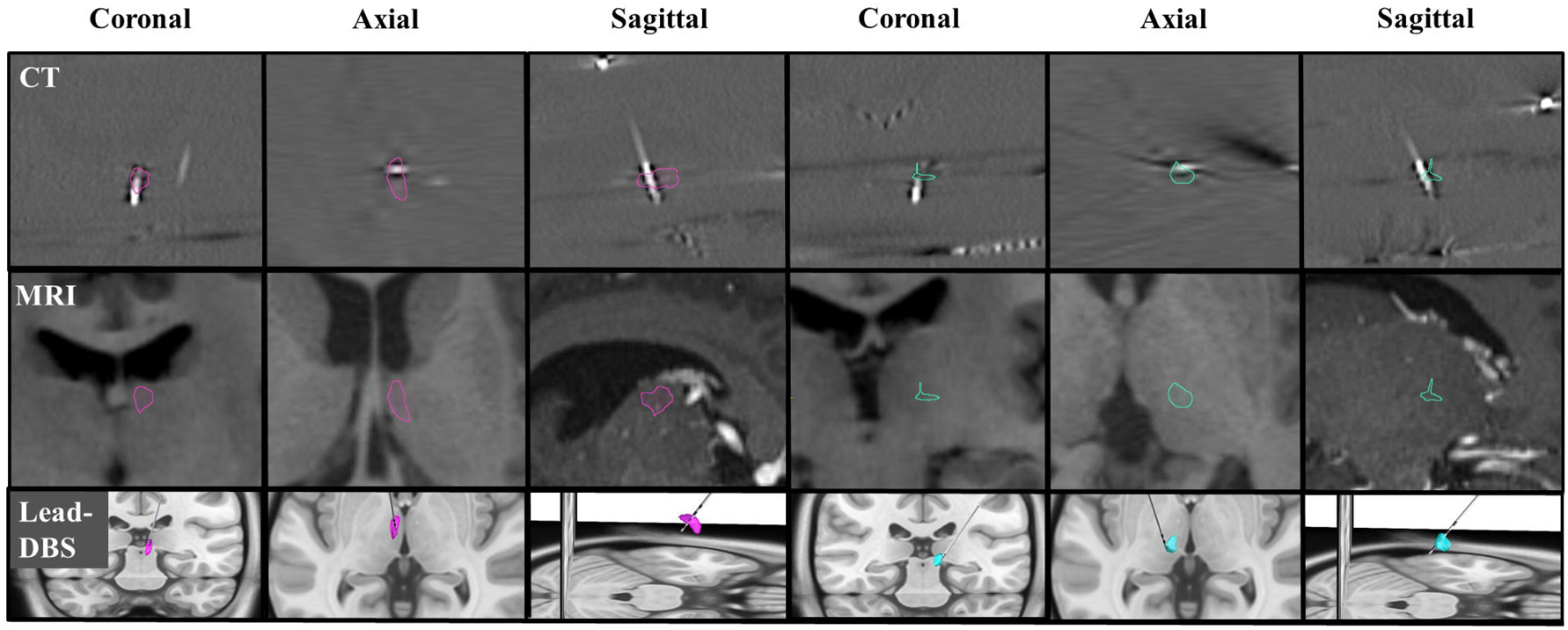
**Thalamic nuclei targeting** Post-op CT (top row), Pre-op MRI (middle row), and Lead-DBS (bottom row) reconstruction is shown to demonstrate verification of placement of the SEEG electrode into the anterior nucleus of the thalamus (pink) and centromedian nucleus of the thalamus (teal) in coronal, axial, and sagittal planes respectively.

### 2.4) Visual analysis of ictal EEG activity

The SEEG data were reviewed by two board-certified pediatric epileptologists (JM, HN) using a Nihon Kohden clinical review station, and the times of cortical and thalamic seizure onsets were based on visualization of persistent rhythmic waveforms on EEG prior to the onset of habitual seizure behaviors that were not otherwise explained by state changes or consistent with prior observed interictal activity (Asano et al., 2009). Clinical seizure onset was determined by observed patient behavior on video review. Seizure activity in cortical and thalamic leads were all marked and time-stamped.

For each patient, we reviewed up to three habitual seizures with the same cortical onset or clinical semiology. For example, if a patient had two different clinical seizure semiologies or two seizures with similar clinical semiology but clearly different electrographic onsets, then these were considered different seizure types, and a total of six seizures were reviewed.

The SOP in the thalamus was determined and categorized as either low voltage fast activity (LVFA), rhythmic spikes, or delta with superimposed LVFA (D-LVFA). Prior studies on adult epilepsy investigating ictal EEG patterns in the thalamus using the similar visual analysis have been published (Burdette et al., 2020, Pizzo et al., 2021). SEEG waveforms were reviewed on Neuroworkbench at sensitivity between 1-7uV, usually, 5-7uV was sufficient to identify ictal onset in the thalamus with a HFF of 70 Hz and LFF of 0.016 Hz.

### 2.5) Quantitative ictal EEG analysis using wavelet transformation

After cortical and clinical onsets were visually marked, ictal SEEG data for the two minutes before and after each seizure at cortical onset were exported in European Data Format (.edf) format. Cortical electrodes used for ictal onset were selected based on where seizure onset was first observed. The ictal thalamic EEG onsets of beta and gamma frequencies were analyzed using EEGLAB2021.1(Delorme and Makeig, 2004) running under Matlab 2021a and in-house developed Matlab code. The EDF data were imported to EEGLAB and downsampled to 200 Hz. The continuous data recorded at each electrode location was decomposed using a Morse wavelet using Matlab function cwt. Logarithmically distributing 141 frequency bins ranged from 0.68 Hz to 86.82 Hz. The obtained scalogram coefficients were converted to log-transformed power using 10xlog_10_ and amplitude. The calculated values within the following frequency bins were averaged to represent conventional EEG frequency ranges: delta (below 4 Hz), theta (4 to 8 Hz), alpha (8 to 13 Hz), beta (13 to 30 Hz), and gamma (30 to 70 Hz). The obtained time-series data were smoothed using a moving window average with a 3-s length. For baseline correction, a 30 second window, from -30 to -60 seconds, prior to the cortical onset was used to calculate mean power across time for subtraction unless severe noise was present during that window. In that case (only 2 cases), custom baseline windows were tailored to avoid artifacts. To show the relationship of cortical and thalamic EEG changes with frequency and the full power spectra, we generated event-related spectral perturbation (ERSP) charts using the standard deviation of the powers in dB during the baseline period (from -60 to -30 s relative to the cortical onset).

### 2.6) Statistical Analysis

The main goal of the analysis was to detect the latency when the signal amplitudes exceeded over two standard deviations (SD), which is equivalent to uncorrected p=0.046 given a Gaussian distribution, from the data during the baseline period. The baseline period was defined as from -60 to -30 s relative to the cortical onset. Thus, we used two standard deviations as an approximate indicator of significance.

For visualization, we calculated a time series of log-transformed EEG power in the result plots, which is suitable for plotting multiple time-series data with relatively large-scale differences.

For EEG power visualization, we also plotted two standard deviation lines calculated from the baseline window for the aid of visual evaluation. Though the two statistics generally agreed, the final results were always adopted from the amplitude data. To detect the onset of pre-onset signal augmentation, the first latency in which the signal amplitude exceeded 2 SD based on the baseline period was counted.

## 3. RESULTS

### 3.1) Cohort characteristics

Nine patients met the inclusion criteria of this study. The age at thalamic recording was 2-25 yo, median age at thalamic recording was 18 yo, with four females, and an average of 79 depth electrodes, and unilateral SEEG AN and CM implant in 8 patients, and bilateral AN in one patient (**Table 1**). Epilepsy types included Tuberous Sclerosis Complex, Rasmussen’s encephalitis, focal cortical dysplasia (FCD), encephalocele and unknown with associated seizure types that included focal motor, focal impaired awareness, focal motor to bilateral tonic-clonic, myoclonic, epileptic spasms, and startle-induced There were no patients excluded.

**Table 1:** Cohort Characteristics.

| Pt No. | Sex | Age at SEEG (yr) | No. of seizures | Epilepsy Type | Etiology | Semiology | MRI lesion/FDG -PET abnormality | Prior surgery | Neocortical Onset | Sampling location | Thalamic Electrode Location | Subsequent RNS | RNS lead location |
| --- | --- | --- | --- | --- | --- | --- | --- | --- | --- | --- | --- | --- | --- |
| 1 | M | 0-5 | >40 | LGS | TSC | Focal tonic and Epileptic spasms | L cortical & subcortical Tub, subependymal nodules along L FTP/L FTP | L laser ablation, CC, L T lobectomy & L O Tub resection | R Fp Tub, L Ant F Tub | Right: F, T, P Tub<br>Left: F Tub | R AN/CM | No | NA |
| 2 | M | 6-10 | >100 | Focal | R ethmoid encephalocele | Focal Motor seizures | R F gliosis/R F | R F pole resection | R Post P | Right: F, T, P<br>Left: F, T, P | R AN/CM | Yes | Cortical |
| 3 | F | 11-15 | 7 | Focal | Unknown | Focal startle induced myoclonic seizures | NL/R T | VNS | L F Op;<br>R F Op | Right: F, T<br>Left: F, T | R AN/CM | Yes | Bithalamic CM |
| 4 | F | 21-30 | 7 | TLE | Unknown | Focal hypermotor | NL/L T | None | L Ant Hip;<br>L Post T | Left: T and Insula | L AN/CM | Yes | Cortical |
| 5 | M | 16-20 | >100 | Multifocal | Unknown | Focal hypermotor | NL/NL | None | R F Op | Right: F, T<br>Left: F | R AN/CM | Approved | NA |
| 6 | F | 16-20 | 10 | Opercular | Rasmussen encephalitis | Focal tonic | L T atrophy & L lateral ventricle dilation/L F | VNS | L Op | Right: T<br>Left: F, T | L AN/CM | Yes | Cortical |
| 7 | F | 21-30 | 2 | TLE | L T FCD | Focal motor | L TPO gliosis/L TPO | L T resection, VNS | L Insula;<br>Inf Post T | Left: F, T | L AN/CM | Yes | Cortico-Thalamic ANT |
| 8 | M | 21-30 | 11 | BiT | Encephalitis & FCD | Focal impaired awareness | L T hyperintense FLAIR Signal/NL | VNS | L Inf T,<br>L Ant Hip; R Op, R Inf T | Right: T<br>Left: F, T | L AN & R AN | Yes | Bithalamic CM |
| 9 | M | 16-20 | 4 | Focal | Unknown | Fip with bilateral tonic-clonic | NL/L TP | None | L O, R O,<br>R AG | Right: T, O, P<br>Left: T, O | L AN/CM | Yes | Cortico-Thalamic CM |

Abbreviations: AN: Anterior nucleus of thalamus; Ant: anterior; biT: bitemporal epilepsy; AG: angular gyrus CC: corpus callosotomy; CM: Centromedian nucleus of thalamus; F: female; F: Frontal; FCD: focal cortical dysplasia; Fip: Focal impaired awareness seizures; Fp: Frontopolar, GTC: generalized tonic-clonic seizure; Hip: Hippocampus; Inf: inferior; L: Left; LGS: Lennox-Gastaut Syndrome; M: male; Mot: motor-premotor epilepsy; NL: Normal; O: Occipital, Op: Operculum; P: Parietal, Post: posterior; R: Right; RNS: responsive neurostimulation; T: Temporal; TLE: Temporal Lobe Epilepsy; TSC: Tuberous Sclerosis Complex; Tub: Tuber; VNS: vagus nerve stimulator.

### 3.2) Accuracy of Thalamic electrode placement

A visual review of electrode location on CT and MRI was done in BrainLab and showed that 94% (17/18) of electrodes had an active contact in or within 1 mm of the nucleus it was intended for. For those leads with no electrode in the intended nucleus, the mean distance from the edge of the electrode to the edge of the thalamic nucleus was 0.4 mm for CM and 1.6 mm for AN. There were no complications, and specifically no intracranial hemorrhage, as a result of placing SEEG electrodes, including thalamic SEEG electrodes.

### 3.3) Seizure characteristics and seizure onset pattern qualitative analysis

On visual assessment of the ictal EEG, there were ipsilateral thalamic EEG changes seen in nearly all seizures at the CM nucleus 97% (29/30), while 86% (31/36) of seizures at the AN showed ictal changes. In the contralateral CM, 100% (4/4) of seizures showed ictal EEG changes, while in the contralateral AN, 71% (5/7) of the seizures showed ictal EEG changes (**Figure 2**). In two patients (1 and 8) with contralateral seizure activity, the AN thalamic leads did not show a clear change in EEG activity.

**Figure 2:**
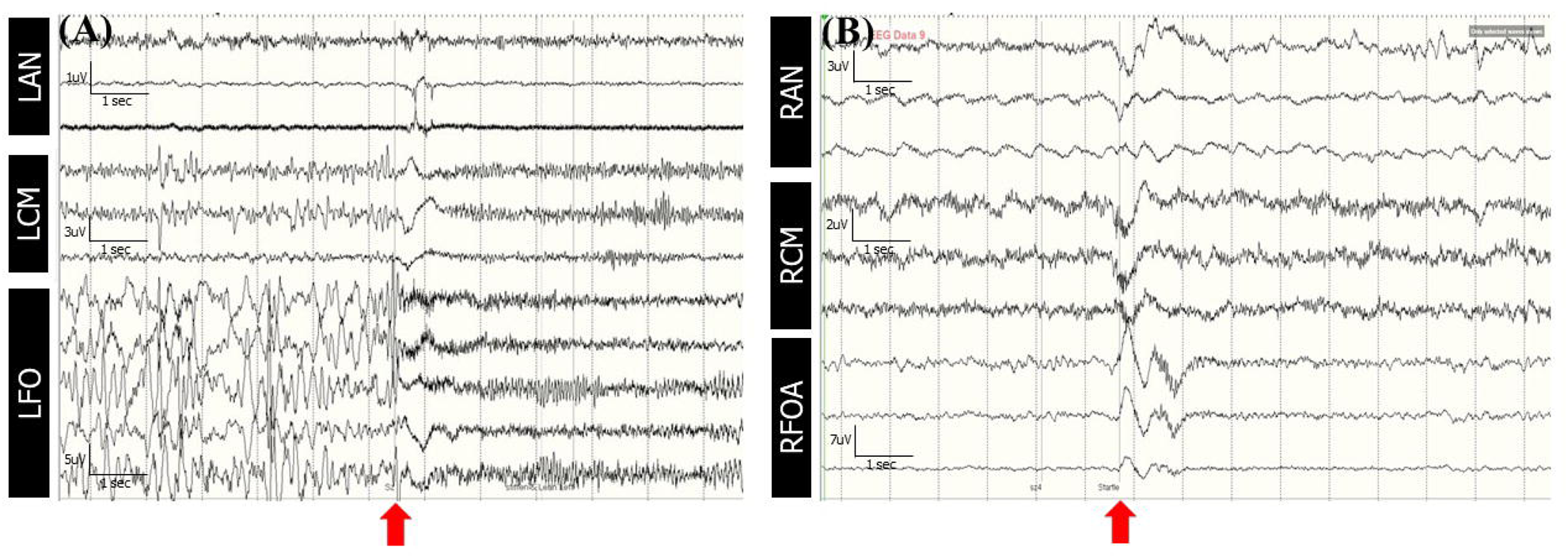
**Stereo EEG** Seizure onset pattern at time of earliest ictal onset in cortex, centromedian and anterior nucleus of the thalamus. Ictal EEG traces with 15 second interval with sensitivity shown to scale. Red arrow indicates seizure onset marked by human experts. (A) Patient 6: rhythmic low voltage fast activity can be seen originating in LFO with rapid spread to CM. (B) Patient 3: High amplitude slow wave in RFOA is followed in RCM by brief period of rhythmic delta with overlying low voltage fast activity. LCM = left centromedian nucleus of thalamus, LAN = left anterior nucleus of thalamus, LFO = left fronto operculum, RCM = right centromedian nucleus of thalamus, RAN = right anterior nucleus of thalamus, RFOA = right fronto operculum anterior.

The SOP recorded most often was delta with superimposed LVFA 46.4% (29/69) with an additional 42.0% (32/69) being LVFA without delta. The next most common patterns seen were rhythmic delta 7.3% (5/69) followed by rhythmic sharps 4.3% (3/69). By location, LVFA was seen in ipsilateral CM 90% (27/29), contralateral CM 100% (4/4), ipsilateral AN 84% (26/31), and contralateral AN 80% (4/5) (Table 2A).

**Table 2:** Qualitative and Quantitative EEG Analysis.

|  | **Ictal EEG**<br>**changes observed** | **SOP**<br>**(most common)** |
| --- | --- | --- |
| Ipsilateral CM | 97% (29/30) | LVFA - 90% (27/29) |
| Contralateral CM | 100% (4/4) | LVFA - 100% (4/4) |
| Ipsilateral AN | 86% (31/36) | LVFA - 84% (26/31) |
| Contralateral AN | 71% (5/7) | LVFA - 80% (4/5) |

**B - Power Analysis (Significance >2SD Threshold)**
|  | **Delta (0-4 Hz)** | **Theta (4-7Hz)** | **Alpha (8-12Hz)** | **Beta (13-30hz)** | **Gamma (31-70Hz)** |
| --- | --- | --- | --- | --- | --- |
| Ipsilateral CM | 88% | 85% | 91% | 94% | 85% |
| Contralateral CM | 67% | 67% | 67% | 83% | 50% |
| Ipsilateral AN | 100% | 97% | 100% | 100% | 91% |
| Contralateral AN | 67% | 83% | 83% | 83% | 100% |

**C - Mean Onset Latency (seconds)**
|  | **Delta (0-4 Hz)** | **Theta (4-7Hz)** | **Alpha (8-12Hz)** | **Beta (13-30hz)** | **Gamma (31-70Hz)** |
| --- | --- | --- | --- | --- | --- |
| Ipsilateral CM | 6.0 [-5.3 to 17.3] | 1.3 [-5.7 to 9.6] | 1.3 [-9.1 to 11.8] | -2.5 [-12.0 to 20.6] | -4.1 [-8.0 to 7.9] |
| Contralateral CM | -4.6 [-13.8 to 4.6] | -7.3 [-15.6 to 1.1] | 6.7 [-9.2 to 22.7] | 13.2 [-53.5 to 32.2] | -11.7 [-39.9 to 5.9] |
| Ipsilateral AN | -0.4 [-9.5 to 8.8] | -5.9 [-13.5 to 1.6] | -0.4 [-9.4 to 8.7] | 0.53 [-6.3 to 15.5] | -0.35 [-8.4 to 17.3] |
| Contralateral AN | -12.7 [-38.0 to 12.4] | -18.0 [-30.7 to -5.4] | -8.5 [-26.2 to 9.0] | -5.3 [-20.1 to 7.1] | 2.0 [-25.3 to 16.2] |

CM: centromedian nucleus of the thalamus; AN: anterior nucleus of the thalamus; LVFA: low voltage fast activity; SD: standard deviation
A) Sensitivity of qualitative ictal EEG changes in the thalamic channels and most commonly observed seizure onset pattern in these locations. B) Quantitative analysis of power change of greater than 2 SD from baseline EEG power in prior 30-60 seconds of Delta to Gamma frequencies. C) Mean onset latency in seconds between power change of 2 SD in cortical onset and power change of 2 SD in CM or AN compared to baseline EEG power in prior 30-60 seconds of Delta to Gamma frequencies.

### 3.4) Quantitative analysis of ictal EEG changes in the thalamic channels

With quantitative broadband EEG analysis of 38 habitual seizures, statistically significant increase, greater than 2 SD, of ictal activity was noted in all frequencies. The greatest percentage of power changes were seen in beta (gamma) activity as noted by significant findings of 93.9% (84.8%), 83.3% (50.0%), 100.0% (90.9%), and 83.3% (100%) at the ipsilateral CM, contralateral CM, ipsilateral AN, and contralateral AN respectively (**Figure 3**, **Table 2B**). Ictal beta and gamma activity emerged around the visual ictal cortical onset: the mean onset latencies (in seconds) of beta activity relative to the cortical onset was -2.5, 13.2, 0.53, and -5.3 at the ipsilateral CM, contralateral CM, ipsilateral AN, and contralateral AN. The mean onset latencies (in seconds) of gamma activity relative to the cortical onset was -4.1, -11.7, -0.35, and 2.0 respectively (**Table 2C**). Broad band analysis was computed on slower frequency bands as well (see details in Table 2).

**Figure 3:**
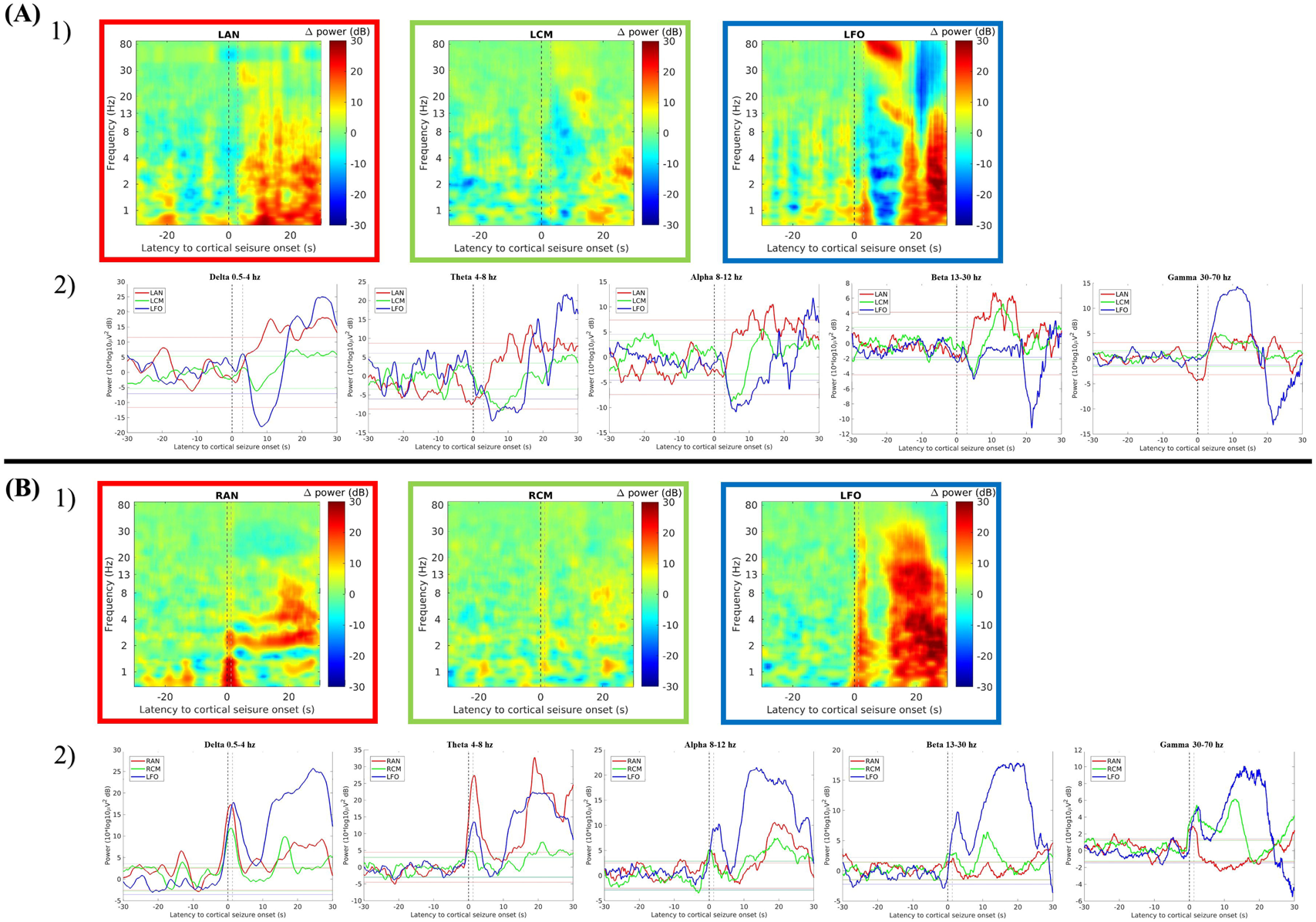
**Quantitative power spectra/frequency analysis** A1 and B1 show ESRP charts showing changes in power across all frequencies by channel LAN, LCM, RAN, RCM, and LFO. (A1) Patient 6: shows significant power change in delta frequencies at the LAN and to less extent LCM with relative increase in power of beta and gamma at LFO. (B1) Patient 3: shows change in delta frequency in RAN at time of seizure as well as significant wide band changes throughout all of cortical channel LFO. A2 and B2 show evaluation of change in power meeting 2SD above baseline power as determined by 30-60 seconds of prior EEG of delta (below 4 Hz), theta (4 to 8 Hz), alpha (8 to 13 Hz), beta (13 to 30 Hz), and gamma (30 to 70 Hz) frequencies in relation to cortical EEG seizure onset (vertical dashed black line) and clinical (behavioral) onset (vertical dashed gray line), and horizontal lines show +/- 2SD relative to the baseline period. (A2) Patient 6: Clear change in power in the LAN and LCM can be seen across all frequencies within 3 seconds of dashed gray line indicating clinical onset. There is also notable LFO sustained increase in Gamma frequencies. (B2) Patient 3: Clear power change can be seen in all frequencies with notable faster frequencies in cortical LFO lead compared to increased Delta and Theta frequencies in RCM and RAN. ESRP = event-related spectral perturbation, LAN = left anterior nucleus of thalamus, LCM = left centromedian nucleus of thalamus, LFO = left fronto operculum, RCM = right centromedian nucleus of thalamus, RAN = right anterior nucleus of thalamus.

### 3.5) RNS experience

A total of seven patients went on to have RNS (two off-label placements due to age, patients 2&3), four with active electrodes, targeting the thalamus (2 bilateral CM, 1 CM & cortical, 1 AN & cortical, 3 cortical). Based on a clear thalamic SOP identified during Phase 2 monitoring we decided to place RNS electrodes for those patients in the AN or CM. Habitual seizures, denoted by magnet swipes, from all four patients with thalamic leads were captured using the detection settings targeting low-voltage fast activities (bandpass detector). Electrical stimulation was then initiated upon ECoG detection of the SOP using this closed-loop system, as demonstrated by patients with cortico-thalamic or bithalamic lead placement (**Figure 4**).

**Figure 4:**
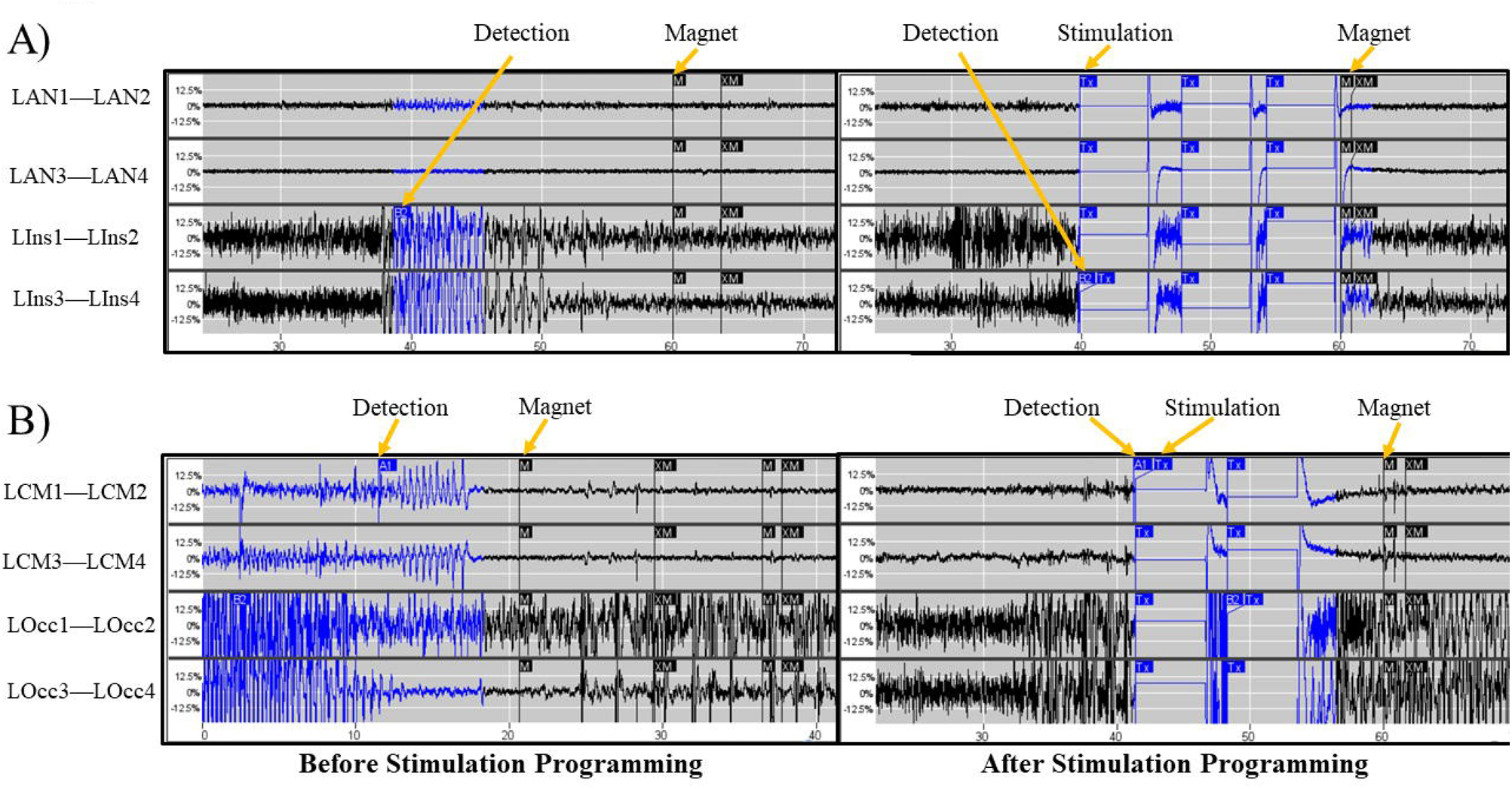
**PDMS ECOG evaluation** ECOG showing two patients with RNS recording in cortico-thalamic (A) and bithalamic (B) arrangement. Detection, Magnet swipe, and Stimulation are marked with yellow arrows. A) Patient 7: demonstrates detection of low voltage fast activity associated with habitual seizure as noted by parental magnet swipe and then subsequent stimulation after RNS stimulator turned on and effective disruption of seizure with stimulation. Ictal thalamic changes were seen almost simultaneously with the cortical onsets. B) Patient 3: demonstrates bithalamic CM ictal onset seen in the RNS recording. With the detection setting adjustment, ictal thalamic EEG changes were detected earlier than the magnet swiping (clinical seizure marking by a caregiver), demonstrating the success of closed-loop stimulations. L: left, R: right, ANT: anterior nucleus, CM: centromedian nucleus, and INS: insular. **– PRINT IN COLOR**

## 4. DISCUSSION

### 4.1) Significance

This is a single-center, retrospective study evaluating the ictal thalamic SEEG recordings from the CM and AN, in nine patients with neocortical focal epilepsy. Using both visual and quantitative analysis, we determined thalamic SOPs and cortico-thalamic seizure onset latency. We found that all seizures captured had ictal activity observed in the thalamus, either in the AN, CM, or both. It most often showed a SOP of LVFA. To our knowledge, our study is the first to compare ictal thalamic EEG changes in the pediatric population with neocortical focal epilepsy at the CM vs. the AN. On expert review, the CM and AN can consistently detect both ipsilateral and contralateral seizures. Our overall findings are robust, having been evaluated with both visual expert interpretation and quantitative analysis.

Our findings imply that recordings from the thalamus can detect habitual seizures in this population and demonstrate preliminary safety and feasibility in routinely targeting thalamic nuclei during SEEG. It further suggests that a closed-loop system such as RNS can feasibly be used to detect and potentially modulate the seizure activity via the thalamus in pediatric patients with (Burdette et al., 2020, Kokkinos et al., 2020, Welch et al., 2021). This provides possible new efficacious neurostimulation options to pediatric patients with neocortical focal epilepsy. Moreover, it further suggests, given the heterogeneous group of patients evaluated, that a wide variety of epilepsies and seizure types, including generalized epilepsies, could be targeted with thalamic neurostimulation that would not otherwise be considered candidates under current device approval.

Study of clinical thalamic recordings date back to at least 1987 when Velasco et. al. (Velasco et al., 1987), inspired by observations of Wilder Penfield (Jasper, 1977), showed that in patients with Lennox-Gastaut Syndrome (LGS) due to multiple etiologies (West syndrome, ischemic injury, FCD, etc) there are distinct recognizable changes in the EEG signal from the CM(Velasco et al., 1991). Furthermore, Velasco et. al. showed that stimulation of the CM in patients with DRE could have significant success. A recent case report by Welch et. al. (Welch et al., 2021) and case series by Beaudreault et. al. (Beaudreault et al., 2022) further demonstrate cases of efficacy in patients with generalized epilepsy receiving neurostimulation at the CM, AN or pulvinar nuclei of the thalamus. Moreover, the largest and most rigorous trial to date, the ESTEL trial, showed that for patients with LGS, neurostimulation with DBS targeting the CM was efficacious even in a short period of follow-up (Dalic et al., 2022). Additional studies have targeted the CM (Fisher et al., 1992, Velasco et al., 2006, Velasco et al., 2000), which has shown potential effectiveness in reducing seizures, especially extratemporal and generalized seizures(Valentin et al., 2013) and patients with LGS(Velasco et al., 2006).

We demonstrated that the AN and CM of the thalamus are both sensitive to cortical seizure onset, as demonstrated by thalamic change seen in 100% of seizures analyzed in this study. There was a non-statistically significant increased sensitivity of ipsilateral CM 97% (29/30) over ipsilateral AN 86% (31/36) with ictal EEG changes being observed. Furthermore, the sensitivity of the CM for contralateral seizure onset of 100% (4/4) vs. contralateral AN 71% (5/7) is intriguing and suggestive of the CM playing a role in bihemispheric cortico-thalamic seizure networks (Lacey et al., 2007, Steriade and Glenn, 1982), whereas this was not seen as consistently from the AN of the thalamus, though given the low number of contralateral seizures in this study this is not a definitive finding. Overall, this preliminarily suggests that only one thalamic lead may be sufficient for bilateral seizure detection for pediatric focal onset seizures.

In this study, the SOP seen in the thalamus was predominately LVFA (88%). This does not seem to differ significantly between the AN and CM. Moreover, the SOP does not change with the type of epilepsy. This is in agreement with prior studies, which have also demonstrated that LVFA is a predominant SOP in the thalamus in extratemporal(Elder et al., 2019, Pizzo et al., 2021) and generalized epilepsies (Velasco et al., 1991). On the contrary, rhythmic spikes patterns were more commonly seen in mesial temporal lobe cases (Pizzo et al., 2021).

The latency between cortical seizure onset and thalamic seizure onset in this study was within 400ms in 95% of cases, based on visual inspection, which is consistent with prior studies (Burdette et al., 2020). We also used quantitative EEG analysis to evaluate the mean time to onset of thalamic ictal activity compared to cortical onset. Importantly, for the quantitative analysis, while we used the broadband frequency data we focused on interpretation of beta and gamma frequencies as this was consistent with the low-voltage fast activity seen most commonly with seizure onset on expert visual analysis, which is the gold standard for evaluation of seizure onset. Interestingly, our findings show that with ipsilateral seizure onset, ictal thalamic EEG activity is detected between at upper and lower bounds of -5.9 seconds to 0.53 seconds (AN) and -4.1 seconds to 6.0 seconds (CM) from the time of cortical ictal EEG activity (Table 2C). The latency ranges for contralateral ictal thalamic activity were more widespread at -18.0 seconds to 2.0 seconds (AN) and -11.7 to 13.2 seconds (CM) (Table 2C). This could indicate less latency variance for the ipsilateral onset and may be associated with anatomical proximity vs. the contralateral side. In rodent models it has been shown that the thalamus is an amplifier of seizure generalization and can show signals with later spread (Brodovskaya et al., 2022). Lag time in the thalamic recordings is also supported by data from prior thalamic recordings that showed patients with LGS had generalized paroxysmal fast activity that was first detected cortically with ∼100ms lag time to it being seen in the CM thalamic nuclei suggesting that thalamic activity is part of the neural network, but not the origin of activity (Dalic et al., 2020, Martin-Lopez et al., 2017). Alternatively, this observation could just be secondary to having fewer seizures for analysis, resulting in the wide range of the confidence interval.

Using quantitative power analysis, we were able to identify changes in all frequencies of >2SD. In beta and gamma frequencies we see significant changes of >2SD for ipsilateral seizures of 94% (CM) – 100% (AN) and 85% (CM) – 91% (AN) respectively (Table 2B). This indicates that ictal EEG changes in amplitude can be detected consistently in most seizures throughout all frequencies. Contralateral seizure detection with > 2SD power change was also noted in all frequencies, though best seen in beta and gamma frequencies at 83% (CM) – 83% (AN) and 50% (CM) – 100% (AN), making the sensitivity not as consistent in contralateral seizures (Table 2B). Our findings suggest that we can feasibly detect habitual seizures by analyzing EEG beta and gamma activity power changes, similar to bandpass settings programmed in the RNS system as has been shown in a prior study(Burdette et al., 2020).

Creating ESRP charts of seizures shows visually the significance of these power changes across all frequencies for the thalamic channels and cortical onset channel (Figure 3). This visualization shows a clear increase in power activity across the frequency spectra with notable changes in delta range frequencies in more than 70% of the thalamic channels, specifically AN, and higher frequency power increases more notable in the cortical channel. This suggests that the response to ictal activity in the thalamus could be slower inhibitory signals in response to cortical activity as opposed to the fast frequency activity associated with ictal onset as discussed previously in temporal lobe epilepsy(Englot et al., 2010) and in evaluation of cortical-thalamic circuits in focal seizures by showing a depressed subcortical arousal response(Motelow et al., 2015). Notably, in most cases, this low frequency power change is not easily seen by visual review and therefore provides another role for the potential importance of quantitative analysis.

Lastly, we demonstrated that we were able to consistently detect a clear SOP associated with habitual seizures (pre-stimulation) in the CM and AN of the thalamus in subjects who were subsequently implanted with RNS.

### 4.2) Limitations & Considerations

Due to the selection of this particular patient population, there were a small number of patients and, therefore, a limited number of seizures analyzed that do not encompass all epilepsy types. Another limitation is that the SEEG placement is not comprehensive and could miss the earliest time point of seizure onset. Furthermore, the determination of the CM, as compared to AN, is challenging given that it does not have clear anatomical landmarks, and a few of these patients have asymmetric brain anatomy making precise lead placement difficult. Moreover, pediatric targeting poses greater challenges given the anatomical brain abnormalities and inability to use standardized cartesian coordinates such as those available in the Schaltenbrand-Bailey atlas. Therefore, some of the thalamic lead placements were not in the intended nuclei affecting the quality of the EEG analysis. Safety of SEEG placement is also an important consideration in this cohort as hemorrhage if to occur in this area would be far more serious than with a cortical electrode and therefore consent should be discussed for all patients. Although we found seizures within 400ms, in most seizures in the CM, it may be that we are underestimating the ability to record seizures in this nucleus given our inability to place SEEG electrodes in the intended position for some. Another consideration is that the computational analysis reviewed was specifically beta and gamma frequencies and did not evaluate high frequency oscillations or low frequency DC shifts.

### 4.2) Future Directions

Regarding future directions, larger and prospective studies are needed to better evaluate the SOP and determine outcomes of seizure reduction by targeting the CM vs. AN with neurostimulation (i.e., RNS). Such studies should have a longer follow-up period to evaluate long-term outcomes of efficacy of RNS and incidence of associated side effects to help guide future programming and optimize effectiveness. Additional studies should further evaluate differences between generalized and multifocal neocortical epilepsy patients, as the networks could vary and affect the target for stimulation and SOP. Furthermore, it would be of interest to characterize broadband information flow in both cortical and thalamic recordings to better understand ictogenesis and propagation patterns of the corticothalamic network during seizures. Using microelectrode recording, it may be possible to further investigate the difference in the function of the AN and CM during seizures at a cellular level (Brodovskaya et al., 2021, Feng et al., 2017).

## 5. CONCLUSION

Our study demonstrates that based on visual and computational analysis, both the centromedian nucleus and anterior nucleus of the thalamus are sensitive in detecting cortical seizure within 400ms of cortical onset and share a common seizure onset pattern of low voltage fast activity, regardless of seizure type or etiology. Our findings suggest that it may be feasible using a closed-loop system in the thalamus, such as RNS, detect and modulate neocortical seizure activity.

## Competing interest statement

All authors have read journal guidelines and there are no declarations of interest.

## Data Availability

All data produced in the present study are available upon reasonable request to the authors

## Acknowledgements

BE is supported by the Epilepsy Foundation Greater Los Angeles. HN is supported by K23NS128318, the Elsie and Isaac Fogelman Endowment, and the UCLA Children’s Discovery and Innovation Institute (CDI) Junior Faculty Career Development Grant (#CDI-SEED-010121; #CDI-TTCF-07012021). HN and RS are supported by the Sudha Neelakantan & Venky Harinarayan Charitable Fund. AF is supported by the Dr. Alfonsina Q. Davies Endowed Chair for Epilepsy Research. MM is supported by NSF 2011716 CRCNS US-Japan Research Proposal: A computational neuroscience approach to skill acquisition and transfer from visuo-haptic VR to the real-world and NINDS 5R01NS047293-16 ‘EEGLAB: Software for Analysis of Human Brain Dynamics’and The Swartz Foundation (Old Field, New York). AD is supported to research abroad by the Uehara Memorial Foundation and SENSHIN Medical Research Foundation. We are grateful to Jimmy C. Nguyen, Andrew Frew, Rajeskar Rajarman, Lekha Rao, and Shaun Hussain for their assistance.

We confirm that we have read the Journal’s position on issues involved in ethical publication and affirm that this report is consistent with those guidelines.

## Competing interest/Conflict of interest statement

All authors have read journal guidelines and there are no declarations of interest.

## Abbreviations

AN: Anterior nucleus of thalamus
CM: Centromedian nucleus of thalamus
DBS: Deep Brain Stimulation
DRE: Drug-resistant epilepsy
ECOG: electrocorticography
ERSP: Event-related spectral perturbation
FCD: focal cortical dysplasia
LAN: Left anterior nucleus of thalamus
LCM: Left centromedian nucleus of thalamus
LFO: Left fronto-opercular
LGS: Lennox-Gastaut Syndrome
LIns: Left insula
LOcc: Left occipital
LVFA: Low voltage fast activity
RAN: Right anterior nucleus of thalamus
RCM: Right centromedian nucleus of thalamus
RFOA: Right fronto-opercular anterior
RNS: responsive neurostimulation
SEEG: stereoelectroencephalography
SD: Standard deviation
SOP: seizure onset pattern
VNS: vagus nerve stimulator.

## Details mentioning exact contribution of each author to work submitted

We certify that all the authors listed made significant contributions to the work and share responsibility and accountability for the results and have approved the final article. MM completed quantitative power/ESRP analysis and latency calculations as well as statistical analysis and figure creation. JM and HN performed EEG review. SA contributed with data collection from RNS. HN provided supervision, data interpretation, study design, and revision of the manuscript. AF, HWP, LR, AB, and NS provided imaging analysis of electrode position. BE wrote primary draft of manuscript, performed literature review and data acquisition. RS and AD provided review of manuscript and study design.

## Competing interest statement

All authors had read and agree they have no competing interest to disclose.

## Notes

### Competing Interest Statement

The authors have declared no competing interest.

### Author Declarations

This is a retrospective single-center study conducted at the University of California Los Angeles. The institutional review board (IRB#18-001599) at UCLA approved the use of human subjects and waived the need for written informed consent, as all testing was deemed clinically relevant for patient care.

